# A comparison of denoising methods in dynamic MRS using pseudo-synthetic data

**DOI:** 10.1101/2021.02.23.21252282

**Authors:** Benjamin C. Rowland, Lasya Sreepada, Alexander P. Lin

## Abstract

**Purpose:** MR spectroscopy of dynamic systems is limited by low signal to noise. Denoising along a series of acquired spectra exploits their temporal correlation to improve the quality of individual spectra, and reduce errors in fitting metabolite peaks. In this study we compare the performance of several denoising methods.

**Methods:** Six different denoising methods were considered: SIFT (Spectral Improvement by Fourier Thresholding), HSVD (Hankel Singular Value Decomposition), spline, wavelet, sliding window and sliding Gaussian. Pseudo-synthetic data was constructed to mimic ^31^Phosphorus spectra from exercising muscle. For each method the optimal tuning parameters were determined for SNRs of 2, 5, 10 and 20 using a Monte Carlo approach. Denoised data from each method was then fitted using the AMARES algorithm and the results compared to the pseudo-synthetic ground truth.

**Results:** All six methods produced improvements in both fitting accuracy and agreement with the ground truth, compared to unprocessed noisy data. The least effective methods, SIFT and HSVD, achieved around 10-20% reduction in RMS error, while the most effective, Spline, reduced RMS error by 70%. The improvement from denoising was typically greater for lower SNR data.

**Conclusions:** Indirect time domain denoising of dynamic MR spectroscopy data can substantially improve subsequent metabolite fitting. Spline-based denoising was found to be the most flexible and effective technique.

## 1 INTRODUCTION

Magnetic Resonance Spectroscopy (MRS) is often used to study time varying metabolic processes in vivo, as it can observe body chemistry non-invasively over a period of time, without perturbing the system it is measuring. Examples of this dynamic MRS include: phosphocreatine (PCr) depletion and recovery in exercising muscle tissue[1], uptake of ^13^C enriched acetate in SAGA (Swift Acetate Glial Assay)[2] and functional MRS[3, 4].

The principal challenge in MRS is to obtain sufficient signal-to-noise ratio (SNR), due to the very low concentration of many of the metabolites of interest. The conventional way to address this issue is to acquire multiple repetitions of the same sequence and average them: as the signal adds coherently and noise incoherently, the SNR increases as the square root of the number of repetitions[5].

In dynamic MRS, however, we are interested in measuring the variation in the system over these consecutive spectra. Combining multiple repetitions into each data point has the effect of degrading the temporal resolution, thus we are forced to choose between characterising spectral features or temporal ones. For example, in a recent publication on muscle spectroscopy at 7 T, Goluch *et al*. used five repetitions and a repetition time (TR) of 6s to obtain a temporal resolution of one data point every 30s[6], whereas for ^13^C MRS at 1.5 T, Sailasuta et al. required 128 repetitions at a TR of 3 s, giving a temporal resolution of 6.4 minutes[2].

The SIFT method (Spectral Improvement by Fourier Thresholding) proposed by Doyle et al. allows improving spectral SNR without affecting the temporal resolution[7]. SIFT works by exploiting the temporal correlation between consecutive acquisitions. For every frequency in the spectrum, the sequence of spectra forms a time series. By reducing the noise along the time series, which we term the “indirect” time domain, the noise within each spectrum is also reduced, leading to an improvement in the quality of fitting. SIFT performs the denoising in the indirect time domain by transforming each time series into the Fourier domain, where the signal is concentrated into a small number of coefficients. Discarding coefficients below a threshold removes the majority of the noise while keeping the large signal coefficients. This process is illustrated in figure 1.

**FIGURE 1.**
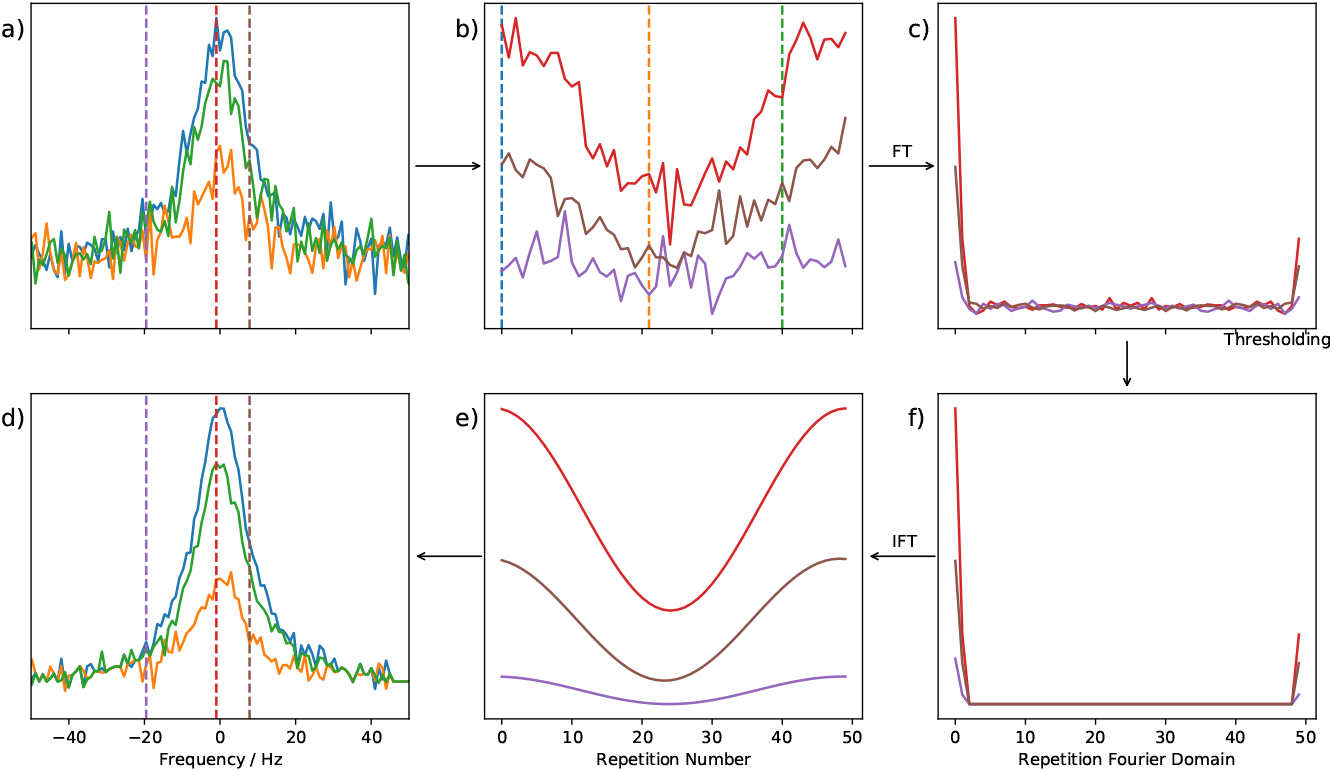
Example of denoising in the indirect time domain with SIFT (Spectral Improvement by Fourier Thresholding). Panel a) shows three simulated spectra of a metabolite peak acquired over a period of time during which it decays and then recovers. The dotted lines indicate three frequencies whose complete evolution over the acquisition period is shown in panel b). Similarly the dotted lines in panel b) show the time points of the spectra in panel a). Panel c) has the Fourier transforms of the time courses in panel b), showing that the signal is concentrated in just a few coefficients. In panel f) a threshold has been applied to keep only the large coefficients, removing most of the noise. Inverse Fourier transform back to the indirect time domain gives the time courses in panel e). Doing this for every frequency we obtain spectra as shown in panel d), which are far less noisy than the original spectra, allowing them to be fitted more accurately during subsequent processing.

The performance of SIFT has been previously demonstrated in dynamic MRS. Rowland *et al*. found that when applied to ^31^P data from exercising muscle, SIFT was able to reduce the standard deviation in the PCr fit by an average of 73%. The improved accuracy of the PCr concentrations, in turn, led to an improved fit of the recovery kinetics, with a reduction in standard deviation for the recovery constant of 38%[1]. However, there are many other denoising techniques that could be applied to achieve this indirect time domain denoising. In this study, we assess for the first time six different denoising methods to see which performs most effectively: SIFT, Hankel singular value decomposition (HSVD)[8], spline fitting, sliding window, Gaussian window, and wavelets. One feature that all denoising algorithms have in common is some form of tuning parameter which determines the aggressiveness with which the noise is removed. If this parameter is set too low, most of the noise is not removed, while if it is too high then some of the signal will be removed along with the noise, distorting the result and possibly leading to misinterpretation of the data. We use a Monte Carlo approach to identify the appropriate tuning parameter for each denoising method for a range of SNRs.

### SIFT

As described above, SIFT works by applying the Fourier transform to the temporal data and then thresholding it. This works as a sparsifying transform, concentrating the signal into a few large Fourier coefficients. The exact choice of threshold (usually measured in noise standard deviations) will affect the performance, with larger values suppressing more noise, but potentially removing signal components. Rowland *et al*. found that using a 2*σ* threshold suppressed around 80% of noise without undue bias to the signal.

### HSVD

The HSVD method is most commonly applied in retrospective water suppression, but is equally applicable in this case. A Hankel matrix is constructed from the temporal data and singular value decomposition (SVD) is used to identify the singular values and associated vectors. These are then truncated to keep only the most significant components, which are used to reconstruct a substantially less noisy signal. The number of residual components, referred to as the rank of the matrix, acts as the tuning parameter for the method.

### Spline

The spline method is technically a smoothing technique - a basis set of cardinal B-splines is fitted to the noisy data. Here there are two tuning parameters available, the order of the splines determines the smoothness of the fitted curve, while the number of splines in the basis set governs how sharply the fit can vary to match rapid changes in the data.

### Sliding Window

Sliding window is the simplest denoising technique, in which each point becomes the average of its neighbors. This is equivalent to convolution with a top hat function. The tuning parameter is the width of the sliding window, i.e. the number of neighbouring points which are averaged to produce each data point.

### Gaussian Window

The Gaussian window is a form of sliding window technique, except that rather than using a top hat window function where each neighbor contributes equally, the points are weighted with a Gaussian function so that those closest to the point being calculated have a greater influence on the final result.

### Wavelets

Wavelet denoising is similar to SIFT in that it begins by converting the signal into a domain with a more sparse representation and then uses a threshold to separate the small noise coefficients from the (mostly) larger signal coefficients. However in this case the wavelet transform is used instead of the Fourier transform. The strength of the wavelet transform is its ability to represent signals which are localised in both time and frequency, however it also adds significant complexity. In this study we used the “Daubechies 8”[9] as the mother wavelet, and a soft thresholding approach.

## 2 METHODS

In order to compare the performance of different denoising methods it is essential to start with a known, noise-free, “ground truth” to which noise is added to form the test data. After applying a given denoising method the result is compared to the original ground truth to see how much noise has been left behind. In this case, our ground truth consists of time courses describing the concentration of each metabolite over a period of repeated acquisitions. From these time courses we can construct idealized artificial FIDs for each acquisition in the series, to which we add Gaussian noise (to both the real and imaginary channels) to create a simulated dataset.

The simulated dataset is then processed using the indirect time domain denoising technique: for each frequency in the spectrum, a time series is constructed from all the repeat acquisitions. Each time series is then denoised with the denoising method of choice and the data points are resorted back into their original spectra. These spectra (which now have reduced noise) are then quantified to obtain metabolite concentration estimates. Finally, the estimated concentrations from the processed noisy data are compared against the ground truth concentrations over the time course, to assess how effective the denoising method has been at recovering the original time course.

### 2.1 Pseudo-synthetic data

When selecting the ground truth concentration time courses, it is important to consider the possibility of introducing bias. For example using a co-sinusoidal model is a simple way to describe a decrease and recovery of a peak, but it innately favours the SIFT method (which decomposes the signal into sinusoids). Similar problems arise with all time courses derived from simple functional forms. In order to test the performance of the method under somewhat more general circumstances, we propose an alternative method, which derives a noise-free time course from experimentally acquired data, which we term *pseudo-synthetic data*.

To create this pseudo-synthetic data, we begin with an experimentally determined metabolite time course, which contains noise, and then remove the noise with the previously described denoising algorithms. Of course, using any individual method to do this would produce exactly the kind of bias that we wish to avoid, as each method will naturally distort the ground truth towards its preferred representation (sinusoids, splines etc.). To mitigate this, we use four separate methods: SIFT, spline, Gaussian window, and SVD (wavelet was not used because it does not produce a noise-free result, only a reduced noise one, while sliding window was not used as its behaviour is similar to Gaussian window), and used a weighted linear combination of the results to form a single time course with elements of each representation.

For a given ground truth, it is easy to see how much it favours the different denoising methods by applying them to it without any added noise: the root mean square error (RMSE) between the original ground truth and the denoised ground truth shows how well the denoising algorithm can represent the original data. To combine our 4 denoised signals into a single ground truth an optimization algorithm was used to determine the optimal combination weights such that applying all denoising method to the combined ground truth would give a similar RMSE between that ground truth and each denoised version. This ensures that there is minimal bias in favour of any of the methods due to the shape of the ground truth, so we are only determining their efficacy in removing noise from signals of this shape.

### 2.2 Exercise Data

Although there are several forms of dynamic MRS which may benefit from temporal denoising, in this study we focus on the widely studied 31P spectra acquired from exercising muscle. The experimental paradigm we follow is as described in [1] for plantar flexion exercises. In brief, the protocol acquired 300 spectra over a 10 minute scan consisting of 3 minutes acquiring a baseline at rest, 2 minutes of exercise and 5 minutes of recovery. The acquisition was a simple pulse acquire sequence with a TR of 2s and 4096 points acquired over a 6KHz spectral width. We created 4 different pseudo-synthetic time courses for phosphocreatine (PCr) and inorganic phosphate (PI) peaks using data acquired from a Siemens Skyra 3T MR system (Siemens Medical Solutions, Erlangen, Germany) and a single channel 10cm 31P tuned transmit/receive surface coil (Mirtech, Boston, USA). The subjects included a young, physically active healthy male, a young healthy female, a middle-aged healthy male and an elderly male suffering from peripheral arterial disease, so that we cover a wide range of different exercise response curves. In each case we normalised the time course so that the baseline period had a mean concentration of 1AU.

Simulated FIDs were constructed from exponential decays using the pseudo-synthetic concentration time courses and the other parameters given in table 1. Gaussian noise was added to both the real and imaginary channels to deliver a desired SNR, defined as the ratio of the FID maximum height to the noise standard deviation. To validate the realism of simulated datasets produced in this manner, we used an experimentally acquired dataset where the subject did not exercise. This case is important as it is the only one where we can know the experimental ground truth, the metabolites are constant in time. Comparing the fitted metabolite concentrations from this experimental dataset against a noise-matched simulated dataset with constant metabolite time courses, we found that their standard deviations agreed to within 2%, indicating that the variability observed over the experimental time course is adequately described by the sources of noise in the synthetic method.

**TABLE 1.**
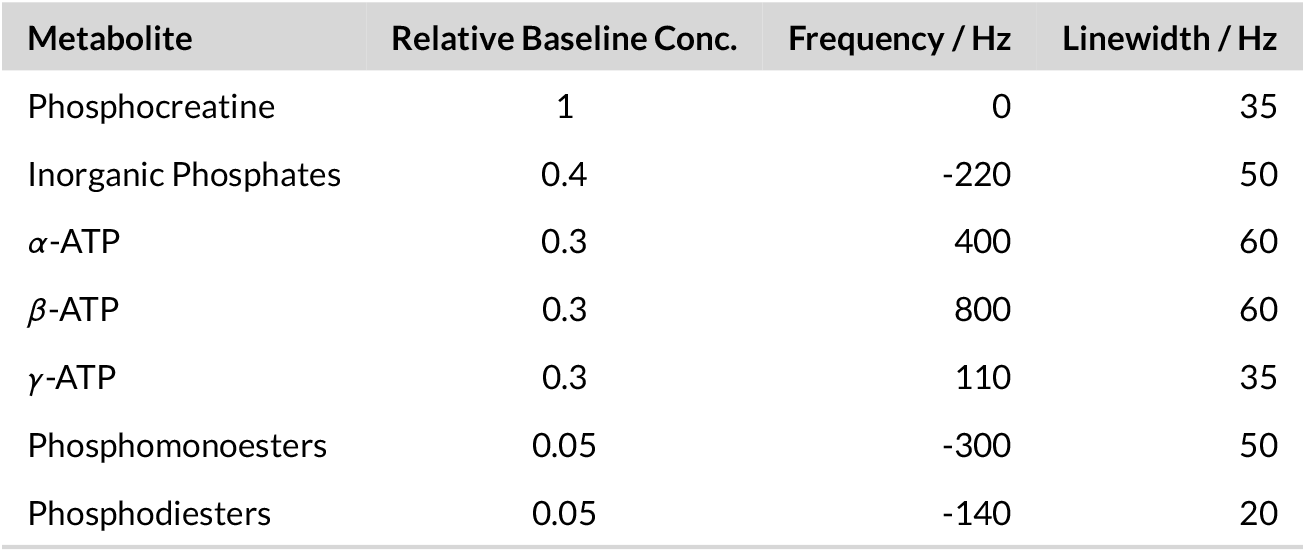
Parameters used in generating pseudo-synthetic data. ATP, Adenosinetriphosphate

| Metabolite | Relative Baseline Conc. | Frequency / Hz | Linewidth / Hz |
| --- | --- | --- | --- |
| Phosphocreatine | 1 | 0 | 35 |
| Inorganic Phosphates | 0.4 | -220 | 50 |
| α-ATP | 0.3 | 400 | 60 |
| β-ATP | 0.3 | 800 | 60 |
| γ-ATP | 0.3 | 110 | 35 |
| Phosphomonoesters | 0.05 | -300 | 50 |
| Phosphodiester | 0.05 | -140 | 20 |

### 2.3 Optimizing the denoising methods

Every denoising method has to strike a balance between removing as much noise as possible and avoiding distortion of the true signal. Each method has at least one tuning parameter, described in table 2 which must be chosen to optimize the performance of the method for the specific problem. Before comparing the different methods it is essential to ensure that they are all being applied optimally. We also wanted to test the performance of the methods at different SNRs to see how sensitive the optimal tuning parameters are to the level of noise. In our experimentally acquired data the mean SNR for an individual spectrum was determined to be 5, therefore we chose to consider SNRs of 2, 5, 10 and 20.

**TABLE 2.** Tuning parameters for the different denoising methods under consideration.

| Method | Tuning Parameter |
| --- | --- |
| SIFT | Threshold in Fourier domain (in noise standard deviations |
| HSVD | Rank of truncated Hankel matrix |
| Spline | Order of spline basis determines smoothness |
|  | Number of splines governs minimum size of features |
| Sliding Window | Number of neighbouring points to be combined |
| Gaussian Window | Number of neighbouring points to be combined |
| Wavelets | Threshold in wavelet domain (in noise standard deviations |

To determine the optimal values of the tuning parameters at each SNR we adopted a Monte Carlo approach. For each method, and for every value of the tuning parameter we were testing, we used our 4 psuedo-synthetic base datasets to generated 100 datasets (25 independent noise signals added to each base simulated signal) which were then processed as described above. All data analysis was performed in Python using the Suspect library[10]. In particular quantification of the metabolite peaks was done using the Suspect implementation of the AMARES singlet fitting algorithm[11]. The RMSE between the estimated metabolite concentrations and the original time course used to generate the pseudo-synthetic data was calculated for both PCr and PI to give the average performance of the method with that tuning parameter. This was then divided by the “no-denoising” RMSE case to get a relative RMSE. Because RMSE naturally decreases with higher SNR, the relative RMSE allows a better comparison of the performance of methods at different levels of SNR. For each technique, we tested out a range of tuning parameter values at each SNR to identify the value giving the optimal performance.

## 3 RESULTS

The complete results of the Monte Carlo analysis are shown in figure 2. As expected, all of the denoising methods are able to offer some reduction in RMSE relative to the raw data, showing that performing denoising can improve quantification accuracy in this time varying data. However, the improvements achieved vary significantly between techniques.

**FIGURE 2.**
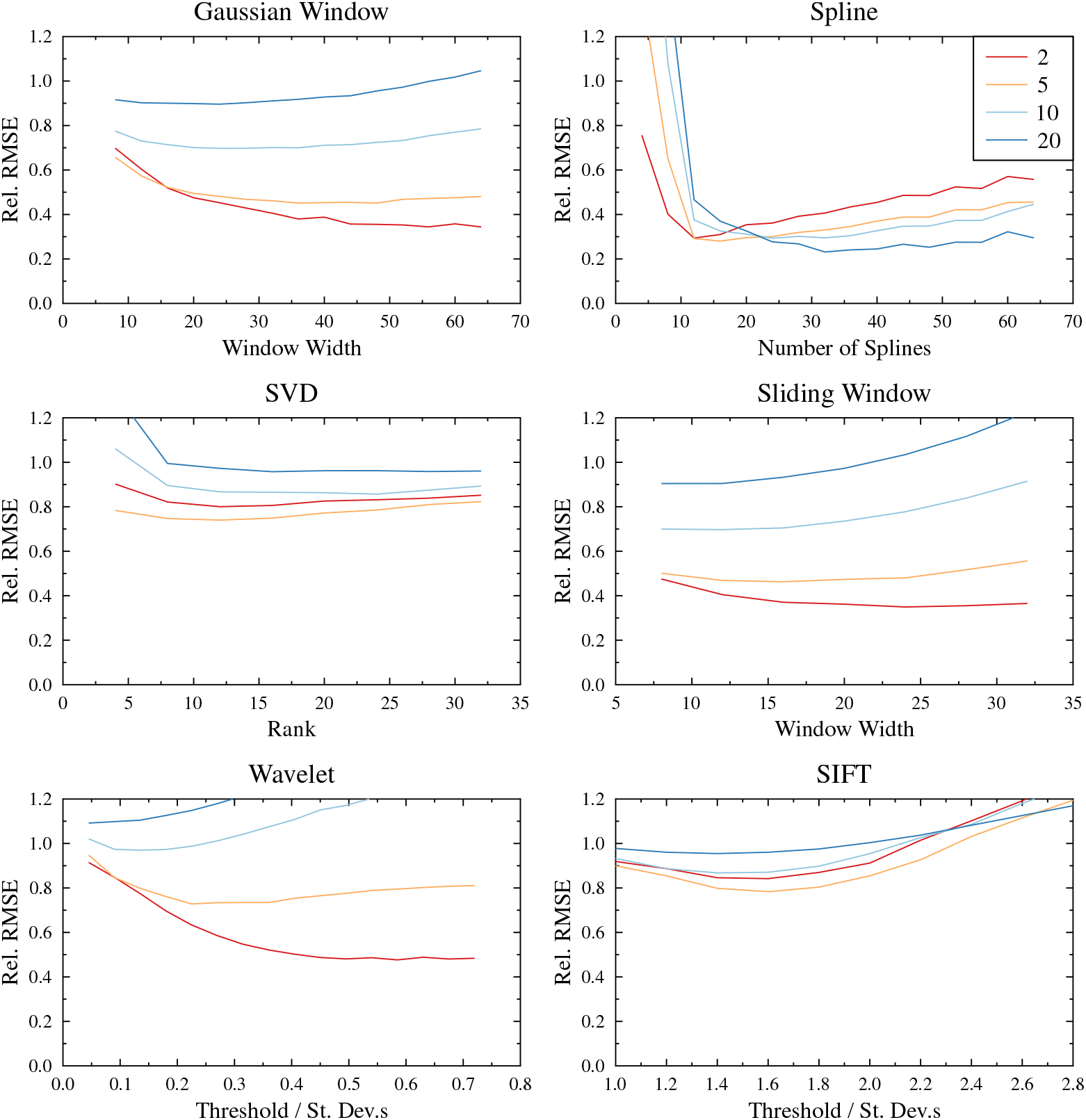
Effect of indirect time domain denoising on quantification of dynamic spectra. Each panel shows the average RMSE between the ground truth and the quantified metabolite timecourse of the denoised pseudo-synthetic data, relative to un-denoised data, as the denoising tuning parameter is varied, for four different levels of SNR.

The SIFT method which has been previously applied to *in vivo* dynamic data actually gives the least reduction in uncertainty. The optimal threshold was found to be 1.6 standard deviations for all levels of noise with a reduction in fitting error of around 20% in the noisier data, but only 5% at SNR 20.

The HSVD method offers slightly improved performance relative to SIFT. A rank of 10 produced the optimal improvement, but above this level the method is very insensitive to changes in the rank.

The wavelet method displayed the greatest variation in performance at different SNRs. For an SNR of 2, using a coefficient threshold above 0.5 standard deviations achieved a reduction in RMSE of more than 50%, but for an SNR of 5 the optimal threshold is only 0.25 standard deviations, while for higher SNRs no improvement at all was possible.

The sliding window and Gaussian window techniques are closely related, so it is unsurprising that their performance is similar. The Gaussian window method is somewhat more insensitive to the effect of the window width on the performance of the method, but the optimal performance is almost exactly the same in both cases, ranging from a 60% drop in RMSE at SNR 2 to a 10% drop at SNR 20.

Finally, the spline technique offers the best performance across the board. The optimal number of splines varies significantly with SNR, from 12 at SNR 2 up to 32 at SNR 20, however the spline method is the only one where the relative improvement in quantification is similar at all levels of noise, with a reduction in RMSE of around 70% at each SNR. An example of one of the pseudo-synthetic datasets before and after spline denoising is shown in figure 3.

**FIGURE 3.**
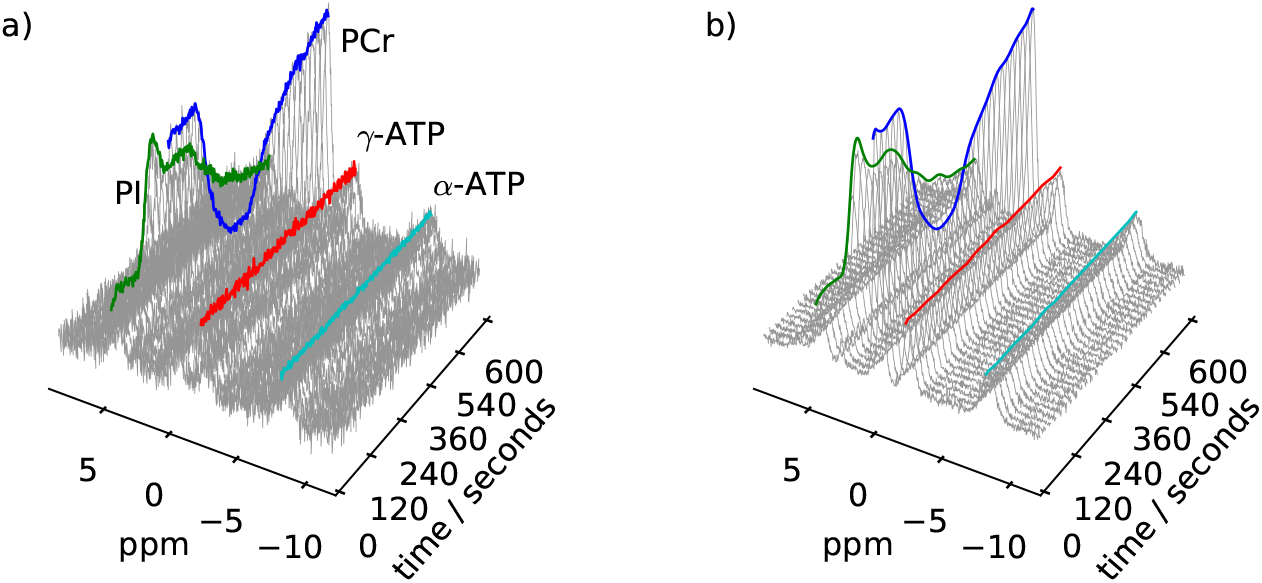
Pseudo-synthetic dataset with SNR 5 before (a) and after (b) applying spline denoising in the indirect tie domain. Also shown are the quantified metabolite concentrations for the metabolites of interest. PCr=Phosphocreatine; PI=Inorganic phosphate; ATP=Adenosinetriphosphate.

A plot of the relative performance of each method is shown in figure 4. For many of the methods shown, the optimal value of the tuning parameter varies between SNRs. However, for *in vivo* datasets it is not always practical to determine the SNR in advance and determine the correct parameter to use. We therefore consider also the globally optimal tuning parameter, which is the value which minimises the mean relative RMSE across all tested noise levels, and evaluate how this single parameter performs against the per-SNR parameters. In every case, we see that the globally optimal parameter very closely matches the performance of the per SNR parameters for SNR5 and SNR10, but generally performs less well for SNR2 and SNR20, although the differences tend to be small even there.

**FIGURE 4.**
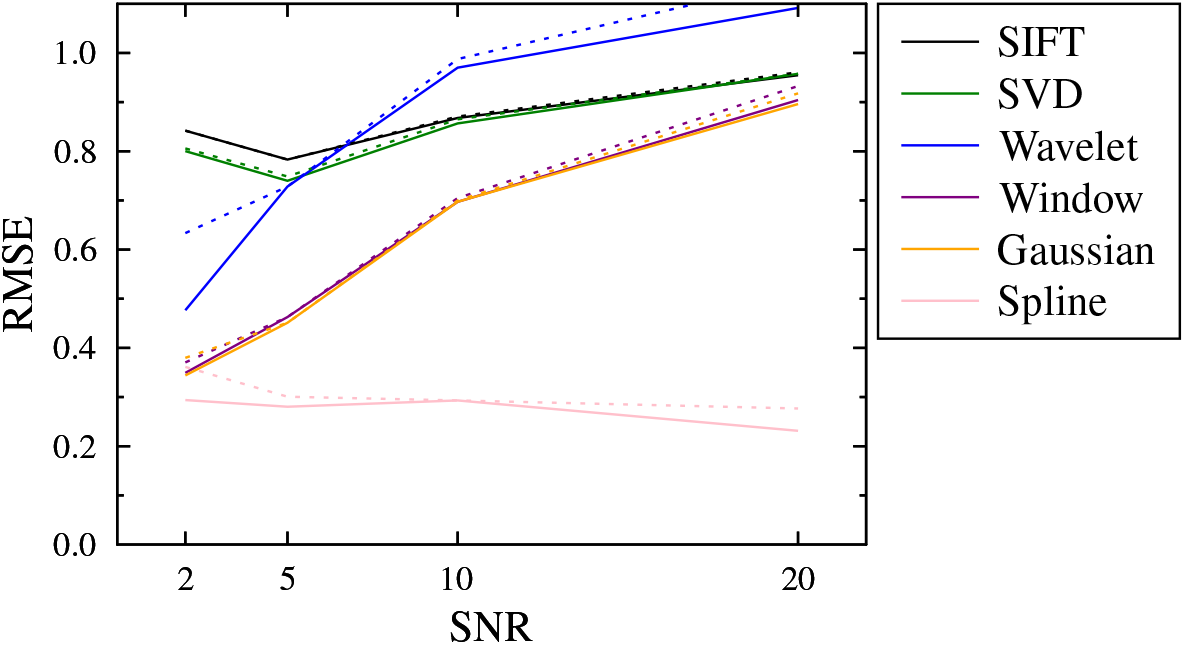
Relative performance of the different denoising methods with optimal tuning parameters. The solid lines show performance with per-SNR tuning parameters, while the dashed line shows performance with a common, globally optimal tuning parameter.

The wavelet method is one of the most popular approaches to denoising in many areas of signal processing, but in this case performed unexpectedly poorly, particularly at higher SNRs where no improvement at all is seen. To understand this behaviour, it is instructive to look at not only the relative RMSE, but also the absolute RMSE across the different SNRs, as shown in figure 5. The wavelet transform attempts to convert the signal into an alternative representation containing a small number of high amplitude signal components and a large number of low amplitude noise components, then uses a thresholding approach to remove the smaller (assumed to be noise) components.

**FIGURE 5.**
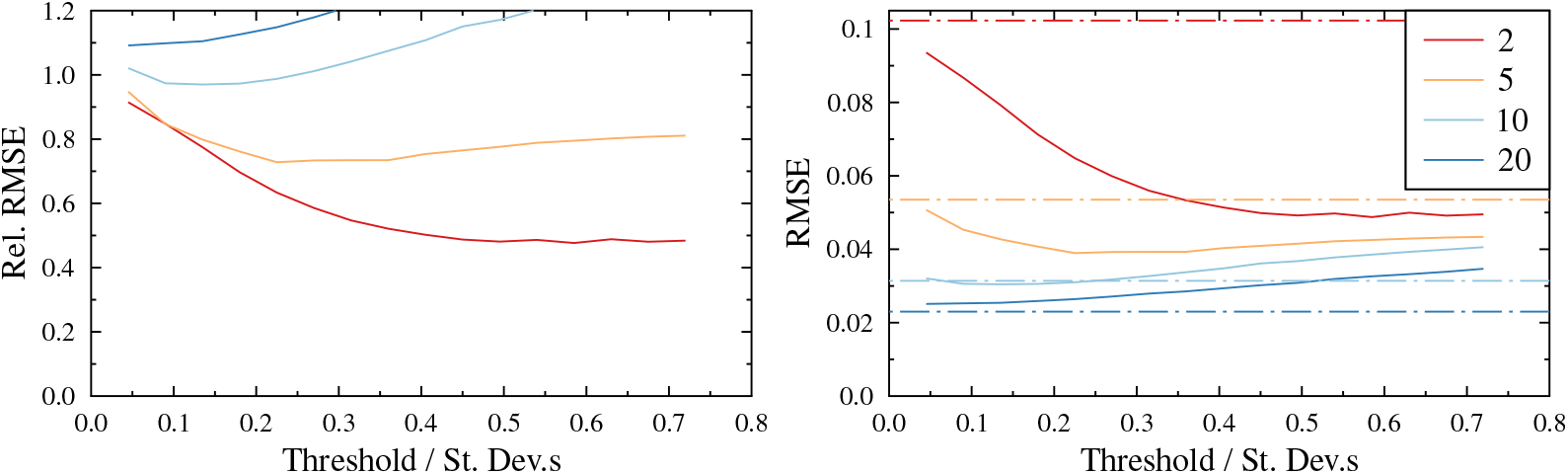
Performance of the wavelet denoising technique shown both relative to raw (un-denoised) data (left) and in absolute performance (right). From the absolute RMSE representation it is clear that as the threshold increases, the wavelet denoising converges to the same representation of the signal irrespective of the SNR. While this is closer to correct than the low SNR raw data, we can see that the spectra are distorted relative to the high SNR raw data.

At very low thresholds, the method reproduces the raw signal almost perfectly, giving no change in RMSE, but as the threshold is increased, we see in figure 5 each SNR converges towards the same value for absolute RMSE. This is because the primary components are the same for all SNRs, so that the same denoised signal is produced in each case. Although this signal appears to be an improvement in relative RMSE at SNR2, at SNR20 we see that the denoised signal is further away from the ground truth than the noisy one: denoising causes an unacceptable deformation of the underlying data, caused by removing too many signal components relative to the noise components. As the results produced at different noise levels are very similar, we can conclude that the same deformation is happening even at SNR2, and for this reason we would recommend against using wavelets even at lower SNRs.

## 4 DISCUSSION

In this work we have explored the application of indirect time domain denoising to dynamic MRS data as an alternative to averaging repetitions to increase spectral SNR and improve quantification. We have compared the performance of six different denoising methods across a range of different SNRs. It is clear that this approach can obtain metabolite concentration estimates which are closer to the underlying true values than those extracted directly from the raw data. However, it is important to bear in mind that removing more noise does not necessarily equate to more accurate results - more aggressive denoising will distort the shape of the data. This is easy to see in the U-shaped curves in figure 2, where the RMSE metric increases with either too much residual noise, or denoising induced distortion. However, the RMSE metric may only be used when the ground truth is known, by using synthetic data. For in vivo data, where the primary metric is fitting uncertainties or Cramer-Rao lower bounds, it is very difficult to know when too much denoising is being applied.

The optimal choice of tuning parameter will vary somewhat with the data being analysed. In our case, the metabolite time courses vary slowly and fairly smoothly, with no sharp transitions. For data which differ substantially from this form, the performance of different techniques, and the optimal values of tuning parameters, may change. It is also important to understand how performance will vary if the selected tuning parameter is not exactly optimal, which may well be the case for experimental data. For our spline data, for example, there is a relatively sharp transition point in the number of splines, below which performance drops off very rapidly, but above which performance remains relatively stable. It makes sense therefore to select a value somewhat above this minimum, so that for unknown data there is no danger of hitting this “hard shoulder” of performance.

Indirect time domain denoising is particularly well suited to muscle and exercise studies where a large number of spectra are acquired over a single activity protocol. Similarly acquisitions of infused labelled molecules with ^13^C or ^1^7O would benefit from applying this kind of denoising in order to get either improved temporal resolution or improved SNR[12]. Other functional paradigms, for example in visual stimulation, cognitive testing and pain, tend to make use of repeated task blocks or events, with fewer spectra acquired per event. In this case averaging over the repeated blocks should be preferred, with denoising applied only within a single repeat unit of the experiment.

The other novel feature described here is the use of pseudo-synthetic data to act as a ground truth in optimization studies. The need for a noise-free ground truth which underlies the noisy signal being processed rules out the use of experimentally acquired data, but artificially constructed signals are often not particularly representative of the real world cases they seek to emulate, containing only the features believed to be of interest in the particular case. Pseudo-synthetic data, derived by removing noise from experimental data, is more likely to retain some of the subtle characteristics that would not be introduced by using purely synthetic data. However, it is important to remember that the denoising method chosen will also introduce some bias into the resulting data, for which reason we advocate using multiple denoising methods and compiling an average.

## 5 CONCLUSION

This study compares for the first time six different methods of indirect time domain denoising for improving SNR and fit reliability in time varying spectroscopy, without affecting the temporal resolution. All the methods tested offered improvement over the unprocessed data, with the exception of the wavelet method. Spline fitting gave the strongest performance with a reduction in noise of more than 70% across a range of SNRs between 2 and 20. Optimum results are achieved by tuning the denoising methods to the SNR of the data being considered, but the use of a globally optimal parameter causes generally only a minor reduction in performance.

## Data Availability

Data is not currently publicly available, interested parties should contact the authors.

## ACKNOWLEDGEMENTS

This work was supported by the Office of the Assistant Secretary of Defense for Health Affairs through the Peer Reviewed Alzheimer’s Research Program under Award No. W81XWH-15-1-0412. Opinions, interpretations, conclusions and recommendations are those of the author and are not necessarily endorsed by the Department of Defense.

## Notes

### Competing Interest Statement

The authors have declared no competing interest.

### Funding Statement

This work was supported by the Office of the Assistant Secretary of Defense for Health Affairs through the Peer Re-viewed Alzheimers Research Program under Award No. W81XWH-15-1-0412. Opinions, interpretations, conclusions and recommendations are those of the author and are not necessarily endorsed by the Department of Defense.

### Author Declarations

The IRB at Brigham and Women's Hospital, Boston approved all human research described.

